# Utility of TaqMan Array Cards for detection of Acute febrile illness etiologies in patients suspected of Viral Hemorrhagic Fever Infections in Uganda

**DOI:** 10.1101/2024.11.20.24317616

**Authors:** GG Akurut, L Nyakarahuka, S Whitmer, D Namanya, K Kilama, S Mulei, J Baluku, A Tumusime, J Kyondo, Julius J Lutwama, Trevor Shoemaker, K Patel, Joel Montgomery, JD Klena, S Balinandi

## Abstract

**Introduction:** Attributing a causative agent to acute febrile illnesses (AFI) remains a challenging diagnostic task for clinical and public health laboratories. The need for developing, validating and implementing a multi-pathogen diagnostic assay able to detect a wide range of AFI-associated pathogens should be prioritized in low-resource settings. The use of the previously developed Acute Febrile Illness TaqMan Array card (AFI-TAC), which detects 26 pathogens in less than 2 hours post-sample nucleic acid extraction, was selected for evaluation in Uganda.

**Methods:** This was a cross-sectional retrospective study in which archived viral hemorrhagic fever (VHF)-negative blood and plasma samples were selected. Samples were previously tested and archived at the Uganda Virus Research Institute (UVRI)-Viral Hemorrhagic Fever (UVRI-VHF) Laboratory from August 2018 to March 2019 as part of the routine surveillance program. The VHF case definition included, temperature of ≥38.0 ºC, additionally with bleeding and any other febrile symptoms. Samples were then tested using an AFI-TAC for 35 potential pathogen targets. Selected positive samples were subsequently assessed using a second, pathogen-specific simplex PCR assay. Epidemiological data were linked to the laboratory findings to assess for risk factors for infection.

**Results:** A total of 82 samples were selected for this study, of which 152 (83.52%) were from Uganda, 22 (12.09%) from the Democratic Republic of the Congo, 7 (3.85 %) from South Sudan, and 1 (0.55%) from Kenya. The median age of the patients was 27 years, with 145 (79.67%) samples from adults >18 years. Seven pathogens were detected with the most prevalent being *Plasmodium* spp. 26.92% (49/182) of which 69.4% (34/49) were *Plasmodium falciparum, Yellow fever* virus 2 (1.10 %), non-typhoidal *Salmonella* 3 (1.65%), *Salmonella enterica serovar* Typhi (1.10%), *Leptospira* spp. 1 (0.54%), *Streptococcus pneumonia* (0.55%), and *Rickettsia* spp. (0.55%). Epidemiological correlation revealed a significant relationship between case fatality (p=0.002) and disease while age, occupation, sex and disease were not significant. Cough was found to be the only clinical symptom associated with being infected with *Plasmodium* (p=0.016).

**Conclusion:** The use of TAC is feasible, readily adoptable diagnostic assay for use in Uganda and will be useful when incorporated into the national testing algorithm for the differential diagnosis of AFI during an outbreak and surveillance.

## Background

Viral hemorrhagic fevers (VHF) are acute febrile illnesses (AFI) characterized by fever with body temperatures ≥ 37.5ºC which may persist for longer than 72 hours. Other symptoms commonly associated with AFI include headache, cough, vomiting, diarrhea, fatigue, and convulsions. Establishing a causative agent based solely on clinical presentation remains a challenge due to the non-specific clinical presentation of AFI, making it difficult to infer a particular symptom to a specific disease. And without laboratory diagnostic testing, treatment is often delayed or ineffective (Maze *et al*., 2018). This is especially important in countries that have experienced multiple, and often simultaneous outbreaks of diseases caused by VHFs such as filoviruses, Rift Valley Fever virus (RVFV), and /or Crimean Congo Hemorrhagic Fever virus (CCHFV) that are capable of spreading rapidly through human and animal populations. Yet, these pathogens are of significant public health concern and their detection and control is important for global health security and socio-economic stability(Balinandi *et al*., 2024, Shoemaker *et al*., 2013). In Uganda, because malaria caused by *Plasmodium falciparum* is a major cause of AFI, many patients with fever are placed on antimalarial prophylaxis in the absence of laboratory diagnostic testing (Ghai *et al*., 2016) which has resulted in several cases of antimalarial resistance (Agaba *et al*., 2024), or generally what is called Antimicrobial resistance (Mambula *et al*., 2023)in Uganda. Moreover, the approach of symptomatic treatment in hospitals has been associated with the under-diagnosis of other AFIs known to exist in Uganda that include brucellosis, leptospirosis, rickettsia, chikungunya, Dengue fever and orthopoxvirus infection (Kharod *et al*., 2016), as has been revealed in the recently implemented Mortuary Surveillance Program (manuscript in preparation).

Diagnosis of a VHF is highly dependent on the direct detection of the causative virus by culture or the presence of viral nucleic acids in the sample from a suspect case. Although the humoral immune response can be utilized, (Racsa *et al*., 2016) it is not practically used for diagnosis. Traditional methods of clinical microbiology such as isolation which requires BSL-4 lab and convalescent-phase serology are slow and cumbersome, and values from common laboratory parameters like urinalysis, chemistries, and hematology are not unique, although they may guide patient identification. Molecular diagnosis of infectious diseases using nucleic acid amplification techniques (NAAT) such as the polymerase chain reaction (PCR) can be designed to be highly sensitive and specific to detect infectious agents. To increase the efficiency of detection, NAAT methods are being designed to target more than one infectious agent in an assay; however, multiplex methods are often less sensitive compared to the singleplex (Hajia, 2017). Taqman Array Cards (TAC) are microfluidic test cards designed to perform multiple real-time PCR specific for several different pathogens (depending on the TAC configuration) using primers and probes preloaded in to the microwells of a 384 well plate. Between 1-8 samples can be tested in parallel against 12 to 384 assay targets in as little as 10fg of DNA isolated from pure bacterial cultures (Rose *et al*., 2012) using TAC. At the current estimates, the cost of using the TAC is ∼80 USD per sample. When analytical validation using clinical specimens was compared between AFI TAC performance and individual PCR assays, an 88% sensitivity and 99% specificity was observed (Liu *et al*., 2016). Thus, the multi-pathogen detection card is convenient for screening AFIs during outbreaks or routine surveillance, consequently reducing the time taken to ascertain the cause of illness, and therefore prophylaxis. Whereas NGS provides the technical solution for pathogen detection efficiency, conclusive results usually take long to be obtained and it is expensive (Lesho *et al*., 2016). The TAC is designed to avoid amplicon contamination with improved detection efficiency for multiple pathogens.

In Uganda, routine VHF surveillance is conducted year-round at the Uganda Virus Research Institute (UVRI). The majority of samples submitted from symptomatic patients test negative for a VHF, yet due to their acute and fulminant nature of illness, there is a need to identify the causative etiologies these patients suffer from (Balinandi *et al*., 2024). A multi-pathogen detection assay for differential diagnosis of acute febrile illnesses reduces the time taken to detect numerous etiologies which can lead to timely public health interventions. This study utilized the TAC assay to detect alternative etiologies for illnesses in patients whose samples tested negative for VHFs.

## Materials and methods

### Study design and population

This cross-sectional retrospective study utilized 182 archived samples submitted to the VHF laboratory at UVRI from August 2018 to March 2019 as part of the National VHF surveillance program testing. The study was given approval by the Director General of the Ministry of Health, Republic of Uganda.

### Inclusion and exclusion criteria

Patients presenting with high fever, with or without hemorrhagic signs, and that tested negative by real-time PCR using primers specific for VHF viruses including Marburg virus, Ebola viruses (Sudan, Bundibugyo and Zaire), Crimean Congo Hemorrhagic Fever virus (CCHF) and Rift Valley Fever virus (RVF), were included in this study. Six known positive samples (RVFV= 5 and CCFV= 1) were used to verify the TAC assay characteristics. VHF surveillance samples collected between August 2018 and March 2019 were included in this study.

### Sample acquisition and nucleic acid extraction

Study samples were retrieved from storage freezers and thawed at 4ºC overnight. All downstream procedures for RNA extraction used the MagMax RNA isolation kit (ThermoFisher scientific, Carlifornia, USA), according to the manufacturer’s instructions. Briefly, from each sample, 100 μl was removed for RNA extraction using an automated Magmax 15 well bead-based extractor (Applied Biosystems, ThermoFisher scientific, Carlifornia, USA,). Samples were inactivated in a high containment laboratory (HCL) by adding 100 μl of patient blood or nuclease free water (negative control) to 400 μl of lysis buffer to which 2 μl of carrier RNA had been added. This was followed by 10 minutes of incubation before transfer of inactivated samples into sterile cryogenic vials and removal from the HCL to a BSL-2 low-containment laboratory for nucleic acid extraction. Inactivated samples were extracted according to the manufacturer’s instructions and eluted in 90μl of elution buffer. The eluates were transferred to sterile labelled tubes and stored at L 20 °C or kept at 4 ºC for immediate analysis.

### Taqman Array Card preparation and analysis

Taqman Array cards first described by (Liu *et al*., 2016), detecting 26 AFI associated etiologies were used. Each TAC tested a total of six patient samples, a positive and a negative control. Mastermix was prepared to include 1x Ag-Path-ID 2X rt PCR buffer and the enzyme mix (Applied Biosystems ThermoFisher scientific, Carlifornia, USA). Into each microfuge tube, 80 μl of the prepared master mix and 20 μl of the RNA were added. This was followed by brief vortexing to mix them and then a short spin.

TAC cards were equilibrated to ambient temperature in their original packages prior to use. 100 μl of each sample mix was added into the inlet port of each channel, the card centrifuged at 1,200 rpm for 2 min, the ports sealed and removed following the manufacturer’s instructions. Amplification was performed on a VIIA7 real-time PCR system (Life Technologies) using the cycling conditions as follows 20 min 45 °C, 10 min 95 °C, followed by 40 cycles of 15 sec 95 °C, 1 min 60 °C. Samples with a cycle threshold (Ct) fluorescent signal of ≤35 were considered positive for the detected agent.

The data was organized in excel and analyzed in R-studio (MA, Boston, USA). Univariate analysis was done to determine demographic characteristics and the frequency of disease occurrence among the different demographic characteristics. A generalized linear model (glm) was developed to determine the general influence of occupation, age and gender on the acquisition of a disease; this was done to attain estimates (odds ratios), and F-statistics all measured at a 95% level of significance. Logistic regression modelling was done to determine the relationship between clinical symptoms and *Plasmodium* infections and the model was validated using the Akaike information criterion (AIC) value.

## Results

### Socio-demographic characteristics of study participants

A total of 182 samples collected from August 2018 to March 2019 met the inclusion criteria. Of the 182 samples, 64.29% were from males, the median age of the patients was 27 years (range 0-80 years) and 79.67% were adults. The geographical distribution of samples selected under this study was as follows; Uganda (n=149; 81.7%)), Democratic Republic of Congo (n= 21; 11.8%), South Sudan (n=7; 3.76%), Kenya (n=1; 0.54%), and n=2; (2.15%) having no country of residence recorded on the case report form. From Uganda, the regions were divided into three, West (n=106; 1%), Central (n=27; 18%) and North (n=16; 11%), (refer to Table 1).

**Table 1:**
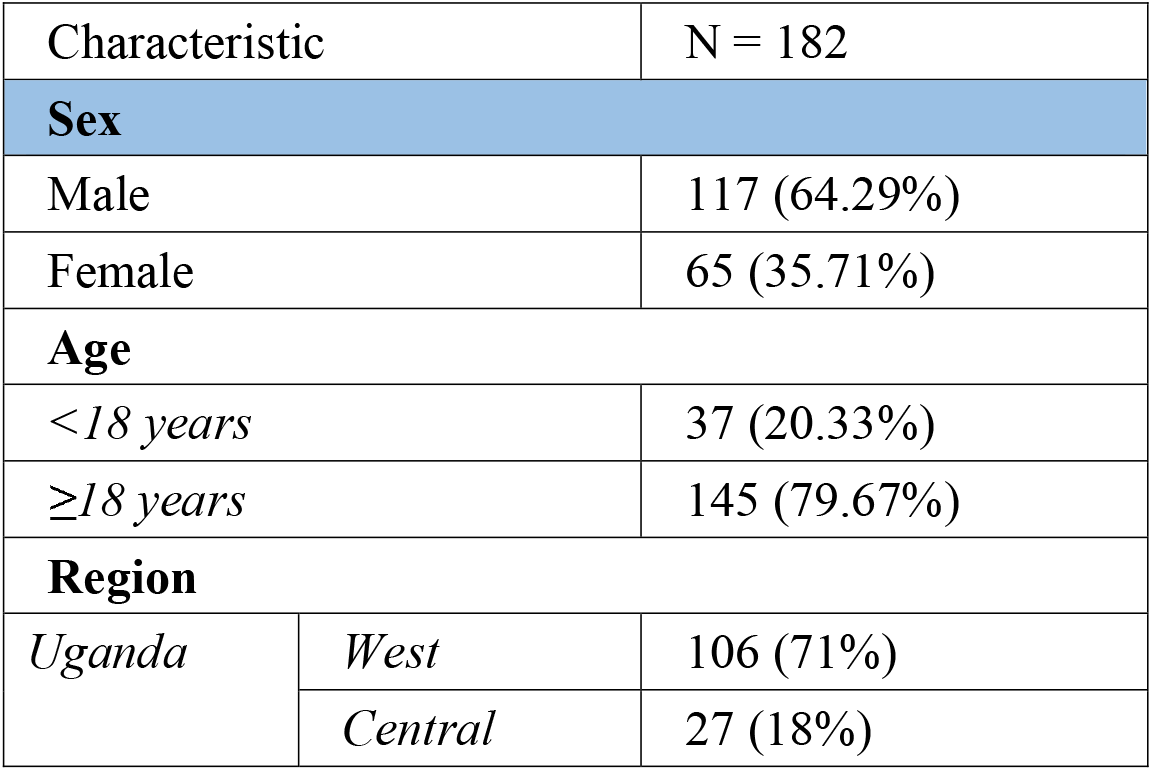
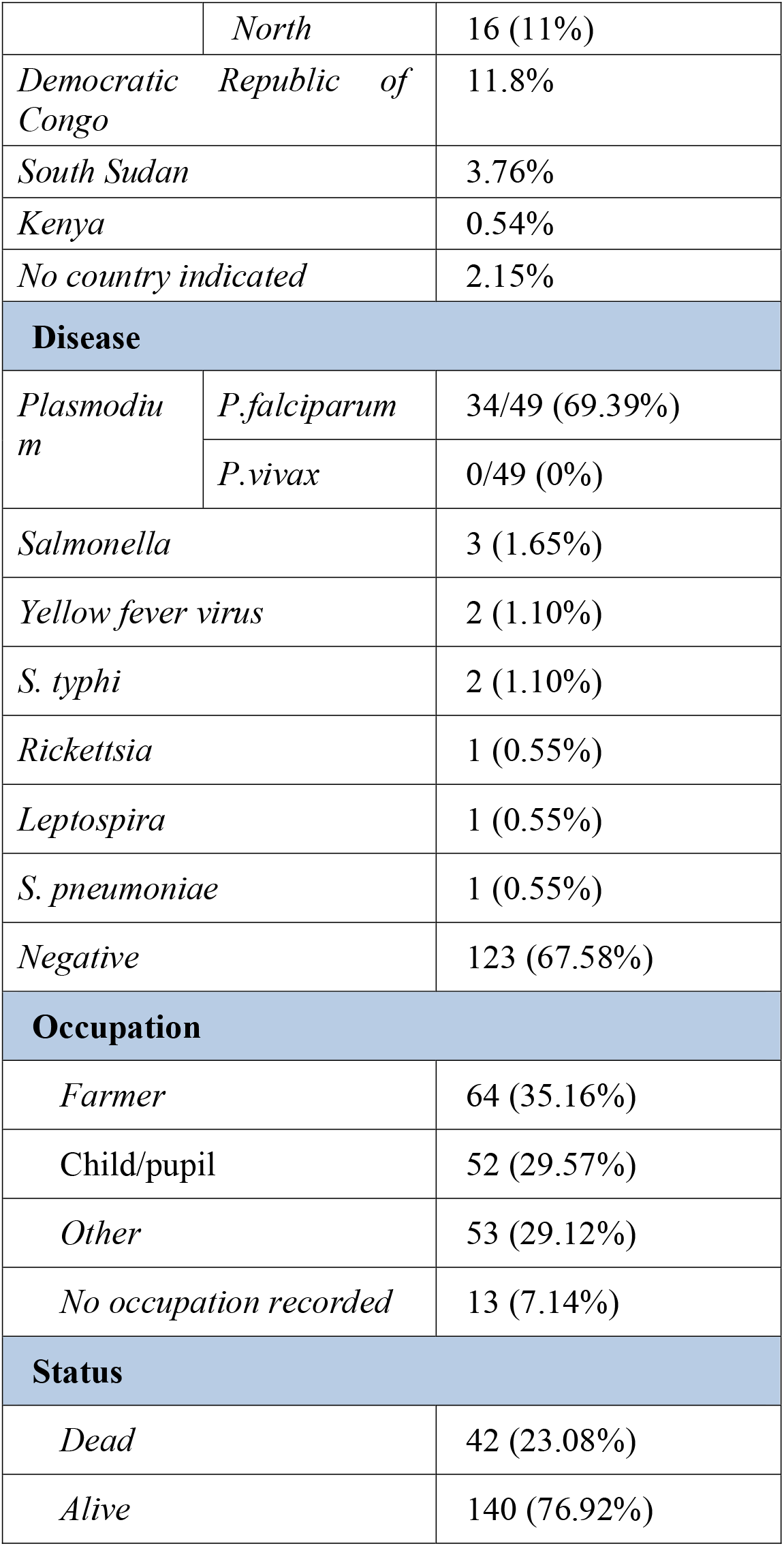
Univariate analysis of disease outcome and socio-demographic parameters and regions within Uganda.

| Characteristic |  | N = 182 |
| --- | --- | --- |
| **Sex** |  |  |
| Male |  | 117 (64.29%) |
| Female |  | 65 (35.71%) |
| **Age** |  |  |
| <18 years |  | 37 (20.33%) |
| ≥18 years |  | 145 (79.67%) |
| **Region** |  |  |
| Uganda | West | 106 (71%) |
|  | Central | 27 (18%) |
|  | *North* | 16 (11%) |
| *Democratic Republic of Congo* |  | 11.8% |
| *South Sudan* |  | 3.76% |
| *Kenya* |  | 0.54% |
| *No country indicated* |  | 2.15% |
| **Disease** |  |  |
| *Plasmodium* | *P.falciparum* | 34/49 (69.39%) |
|  | *P.vivax* | 0/49 (0%) |
| *Salmonella* |  | 3 (1.65%) |
| *Yellow fever virus* |  | 2 (1.10%) |
| *S. typhi* |  | 2 (1.10%) |
| *Rickettsia* |  | 1 (0.55%) |
| *Leptospira* |  | 1 (0.55%) |
| *S. pneumoniae* |  | 1 (0.55%) |
| *Negative* |  | 123 (67.58%) |
| **Occupation** |  |  |
| *Farmer* |  | 64 (35.16%) |
| *Child/pupil* |  | 52 (29.57%) |
| *Other* |  | 53 (29.12%) |
| *No occupation recorded* |  | 13 (7.14%) |
| **Status** |  |  |
| *Dead* |  | 42 (23.08%) |
| *Alive* |  | 140 (76.92%) |

### Pathogens detected by TAC

TAC detected 7 pathogens targets in (32%) samples (Table 1). The major etiologies detected were *Plasmodium* spp., 26.9% (49/182), particularly 69% (34/49) were determined to be *Plasmodium falciparum* species. Other etiologies detected include; *Salmonella* spp. (n=3; 1.65%), while Yellow Fever virus (n=2; 1.1%) and *S*.*typhi* (n=2; 1.1%) were both detected in two samples. *Streptococcus pneumonia, Leptospira* and *Rickettsia* (0.55%) were each detected in a single sample.

There was a statistical significance between case fatality and having disease (P value = 0.002 95% CI). That Case fatality rate of Leptospirosis, Rickettsia, *Salmonella* Typhi, *Yellow fever* and *S. pneumoniae* was 100%, while it was 16% and 33% for plasmodium and salmonella infections, respectively (Table 2). Samples previously positive for individual real-time PCR (IRTP) RVF (n=5) and CCHF (n=1) were included for assay verification and there was 100% agreement as they also tested positive by TAC. The samples that tested positive for Yellow Fever virus by TAC were found positive by individual real time PCR. There was no statistical significance at 95% confidence that any of the clinical characteristics presented were associated with Plasmodium infection besides cough (p value = 0.016 95% CI) (refer to Table 3).

**Table 2:** Percentage frequency of diseased individuals among different demographic parameters.

| Category | n | Overall<br>N=182 | Lpt<br>n=1 | Plas<br>m | Rk<br>n=1 | Sal<br>n=3 | S.p<br>n= 1 | S.th<br>n=2 | YF<br>n=2 | p-value |
| --- | --- | --- | --- | --- | --- | --- | --- | --- | --- | --- |
|  |  |  |  | **n=49** |  |  |  |  |  |  |
| **Sex** | 182 |  |  |  |  |  |  |  |  | 0.9 |
| *F* |  | 36% | 0% | 43% | 0% | 33% | 0% | 50% | 0% |  |
| *M* |  | 64% | 100% | 57% | 100% | 67% | 100% | 50% | 100% |  |
| **Age** | 182 |  |  |  |  |  |  |  |  | 0.9 |
| *<18* |  | 20% | 0% | 27% | 0% | 0% | 0% | 0% | 0% |  |
| *≥18* |  | 80% | 100% | 73% | 100% | 100% | 100% | 100% | 100% |  |
| **Occupation** | 169 |  |  |  |  |  |  |  |  | 0.3 |
| *Farmer* |  | 38% | 100% | 38% | 0% | 0% | 100% | 0% | 50% |  |
| *Child/pupil* |  | 31% | 0% | 40% | 0% | 33% | 0% | 0% | 0% |  |
| *Other* |  | 31% | 0% | 22% | 100% | 67% | 0% | 100% | 50% |  |
| **Status** | 182 |  |  |  |  |  |  |  |  | **0.002** |
| *Dead* |  | 23% | 0% | 16% | 100% | 33% | 100% | 100% | 100% |  |
| *Alive* |  | 77% | 100% | 84% | 0% | 67% | 0% | 0% | 0% |  |
| **Region** | 141 |  |  |  |  |  |  |  |  | 0.4 |
| *North* |  | 11% | 0% | 19% | 0% | 0% | 100% | 0% | 0% |  |
| *West* |  | 71% | 100% | 62% | 10% | 67% | 0% | 100% | 0% |  |
| *Central* |  | 18% | 0% | 19% | 0% | 33% | 0% | 0% | 0% |  |

*Disease among categories; sex, age, occupation, status and region presented in percentages.*
*Status; dead or alive is more associated with having the disease (statistically significant).Lpt;*
*Leptospira, Pl; Plasmodium, Rk; Rickettsia, Sl; Salmonella, S.p; Streptococcus pneumonia, S.t;*
*Salmonella typhi, YF; Yellow fever*

**Table 3:** Association between clinical characteristics including abdominal pain, chest pain, muscle pain, headache, cough, sore throat, conjunctivitis and skin rash and presence of Plasmodium infections., cough is the only statistically significant clinical characteristic.

| Characteristic | OR | 95% CI | p-value |
| --- | --- | --- | --- |
| Fever | 2.69 | 0.76, 13.1 | 0.2 |
| Bleeding | 1.27 | 0.50, 3.42 | 0.6 |
| Vomiting | 0.95 | 0.43, 2.10 | 0.9 |
| Diarrhea | 0.71 | 0.32, 1.56 | 0.4 |
| Anorexia | 1.21 | 0.57, 2.61 | 0.6 |
| Abdominal pain | 0.96 | 0.45, 2.08 | >0.9 |
| Chest pain | 2.13 | 0.89, 5.20 | 0.092 |
| Muscle pain | 1.09 | 0.51, 2.30 | 0.8 |
| Headache | 1.33 | 0.62, 2.98 | 0.5 |
| Cough | 0.32 | 0.12, 0.78 | **0.016** |
| Sore throat | 0.81 | 0.26, 2.33 | 0.7 |
| Conjunctivitis | 1.00 | 0.34, 2.71 | >0.9 |
| Skin rash | 1.33 | 0.44, 3.90 | 0.6 |
| <sup>1</sup>OR = Odds Ratio, CI = Confidence Interval |  |  |  |

## DISCUSSION

Multiplex pathogen detection tools like TAC PCR can be valuable tools for diagnosing multiple infections for infectious disease surveillance (Rose *et al*., 2012, Hercik *et al*., 2017). Because these systems are open, they can be custom-designed to investigate possible causes of syndromic disease (e.g. acute febrile illness) or for priority diseases within a country or region and environmental samples testing (Kodani *et al*., 2011).

Prior to this study, after completion of the VHF testing algorithm (using simplex assays for Ebolavirus, Sudan Ebolavirus, Bundibugyo Ebolavirus, Marburg virus, Crimean-Congo Hemorrhagic fever virus and Rift Valley fever virus) at UVRI, no further testing would be performed on a sample. In this study, of the 182 VHF-negative samples tested by TAC, at least one organism was identified in 59 (32.4%) of the samples by AFI-TAC assay, practically meaning that a diagnosis of the infectious cause of disease can be made for 1 of every 3 VHF-negative cases. The most prevalent pathogen was *Plasmodium* spp. interestingly, the two cases of yellow fever virus cases detected by this study triggered an outbreak investigation of this etiology in Masaka district, Central Uganda, by the Ministry of Health and was confirmed by singleplex PCR (https://www.health.go.ug/programs/uganda-national-expanded-program-on-immunisation-unepi/). From this study, Yellow fever virus disease severity and mortality was associated with occupations such as hunting and farming which activities predispose people to mosquito bites. Furthermore, Yellow fever cases were males similar to the outbreak in northern Uganda that found males were more likely than females to acquire the *Yellow fever* virus. This predisposition of males towards YF infection was linked to economic activities undertaken by males around forests (Wamala *et al*., 2012).

Similarly, both non-typhoidal and typhoidal *Salmonella enterica* were detected with non-typhus salmonella (n=3, 1.65%), *Salmonella typhi* (n=2; 1.10%), as well as Leptospira (n=1; 0.54%), and Rickettsia (0.55%). Finally, a single case of *Streptococcus pneumoniae*, was found. A similar study from Tanzania found *Plasmodium* spp. to be the most common pathogen detected (using the identical AFI card while *Leptospira* was only 3% of cases and *Salmonella enterica* and *Rickettsia* were detected in 1% of cases (Hercik *et al*., 2017). A study in Bangladesh found *Salmonella enterica* serovar Typhi (15%), dengue (10%), *Rickettsia*, spp. *Escherichia coli, Pseudomonas* spp., *Leptospira* spp., *Coxiella* spp., *Histoplasma*, and *Toxoplasma*; malaria RDT-positive patients were excluded from this study (Ferdousi *et al*., 2023). Much as *S. pneumonia* is not described as a cause of AFI, it causes high morbidity and mortality especially in children under the age of five *(Zar et al*., 2013)

In this study several pathogens of outbreak potential and concern to the Uganda public health were identified as mentioned above. However, the detection of high consequence pathogens rarely reported in Uganda such as West Nile virus, *Bartonella*, and *Coxiella* was not made. To assure that the assay would have detected a variety of low frequency but high impact pathogens, this study included six RVFV and CCHFV samples (RVFv, n = 5 and CCHFv, n = 1) with a range of Ct values (25-28). All six of these samples were detected as positive for their respective pathogens by TAC with Ct values (26-29). This shows that the performance of TAC is comparable to simplex PCR.

This study found *Plasmodium* infections to be the most prevalent, similar to findings from a study in Kilombero, Tanzania (Hercik *et al*., 2017) where 47% of patients recruited into the study became positive for *Plasmodium*. A study on metagenomic next-generation sequencing of samples from pediatric febrile illness in Tororo, Uganda found the most prevalent AFI etiology to be *Plasmodium falciparum* at 51.1% (Ramesh *et al*., 2019). As much as *P*.*vivax* targets were on the card, 15/49 samples that tested positive for plasmodium still remain unspeciated. The rarity and low prevalence of *P. vivax* in sub-Saharan Africa might owe to the fact that there is a wide range of Duffy negative alleles in the population making them more resistant to *P. vivax* (Dhorda *et al*., 2011) (Howes *et al*., 2011). These samples may be positive for *P*.*malaria* or *P*.*ovale* which have been documented to exist in Uganda (Asua *et al*., 2017). There was no statistical significance at 95% confidence that any of the clinical characteristics presented were associated to plasmodium infection besides cough. Similarly, complicated Malaria has been associated with lung damage, altered pulmonary functions such as Acute respiratory distress with signs such as cough (Cabezón & Hernández-mora, 2016). In this study, all mortalities are associated with an infection which could be associated to delays in diagnosis or misdiagnosis culminating to inadequate patient support.

Some of the limitations of the TAC assay include the need to test 8 samples to avoid waste. In situations where the sample volumes are low plus the necessity to adhere to set turnaround times, the assay becomes expensive. This is because testing less than 6 samples becomes a waste of resources. On the other hand, circumstances of very high sample volumes, testing 6-8 samples may be a limitation to meeting the turnaround time. The products from the PCR cannot be used for any downstream analysis like sequencing. In addition, during development, a single annealing temperature is needed therefore the need for optimization of individual assays for efficiency (Heaney *et al*., 2015).

## Recommandations

From this study, it is eminent to include the TAC card testing as routine for all VHF negative testing done in country. The unspeciated plasmodium positive samples need to be tested for other *Plasmodium* species such as *P*.*malariae* and *P*.*ovale*. Based on the commonly circulating AFI pathogens in Uganda,, the AFI card needs further be customization to include etiologies such as; COVID-19, monkey pox virus, and measles with clear guidance from the Ministry of Health. In addition, a TAC assay targeting travelers can be designed for pathogens that are not endemic to Uganda like *Lassa fever virus*, and *Nipah virus* disease that could otherwise be imported by international travelers from countries where these etiologies are known to exist. While the strategy of duplication of pathogen targets reduces the number of pathogens being detected thus making the assay even more expensive, it could increase the efficiency of the assay.

## Conclusion

Although the TAC cards are expensive, costing between 350 to 500 USD per card, for the number of pathogens tested and the number of samples that can be tested per card, strategic use of the cards could be a cost-effective way to perform passive surveillance for a range of pathogens that are of public health importance to Uganda. The TAC system provides flexibility of testing for multiple pathogen targets within a short period of time and without the need to purchase and maintain multiple kits, positive controls and reference material. Coupled with confirmatory testing at the appropriate reference laboratory for presumptive-positive samples identified using TAC, the system could be effective at more quickly recognizing and responding to outbreaks. Compared to singlex procedures. This study has therefore proved that the AFI-TAC testing modality is of great value and can be adopted to Uganda for AFI surveillance and outbreak investigations.

## Data Availability

All data produced in the present work are contained in the manuscript

## Ethical approval

The study was approved by the office of the Director General of Health services in the Ministry of Health Uganda.

## Acknowledgements

I acknowledge the centers for Disease control and prevention (CDC) who funded this work through the Viral Hemorrhagic Fevers surveillance program at the Uganda Virus Research Institute.

